# Sequencing from leftover material of stored rapid Ag Tests (Ag-RDTs): a gamechanger in genomic surveillance?

**DOI:** 10.1101/2022.06.23.22276745

**Authors:** Annabel Rector, Mandy Bloemen, Gilberte Schiettekatte, Piet Maes, Marc Van Ranst, Elke Wollants

**Author notes:** Corresponding author; Herestraat 49 box 1040, BE3000 Leuven, Belgium.

## Abstract

Archived lateral flow antigen-detection rapid diagnostic tests (Ag-RDTs), used in the diagnosis of COVID-19, were successfully used to extract viral nucleic acids for subsequent RT-qPCR and sequencing by Sanger or Nanopore whole genome sequencing (WGS). The method was successfully applied with different brands of SARS-CoV-2 Ag-RDTs, but also with Ag-RDTs for detection of influenza, rotavirus and adenovirus 40/41. The buffer used in the Ag-RDT had an important influence on the RNA yield from the test stripand the efficiency of subsequent sequencing. Our finding that the test strip in rapid Ag tests is suited to preserve viral genomic material, even for months at room temperature, and therefore can serve as source material for genetic characterization, could improve global coverage of genomic surveillance for SARS-CoV-2 as well as for other viruses.

## INTRODUCTION

With over half a billion cases and more than 6 million reported COVID-19 deaths since it’s emergence in December 2019 [1], the COVID-19 pandemic caused by SARS-CoV-2 has brought on major challenges to health care systems and authorities worldwide. The availability of effective and safe vaccines is without question an important element in our way out of this crisis. Nevertheless, the current COVID-19 vaccines are not sufficient to prevent spread and circulation of the virus, even in highly vaccinated populations [2], and are regretfully not accessible for the entire global population [3]. Therefore, epidemiological surveillance through timely and adequate diagnostic testing for SARS-CoV-2 to guide preventive measures for the control of COVID-19 will remain vital in the battle against the virus.

Currently, two types of tests are being used in the diagnosis of SARS-CoV-2 infection. Molecular assays based on the detection of viral RNA through a nucleic acid amplification test (NAAT), such as real-time reverse transcription quantitative PCR (RT-qPCR), are highly sensitive and specific, but in most cases require expensive laboratory facilities and trained technicians, making them less suited for fast scaling-up. An alternative is the detection of viral antigens through immunodiagnostic techniques such as lateral flow antigen-detection rapid diagnostic tests (Ag-RDTs) that can be visually read or processed and read by an instrument. These Ag-RDTs can be performed outside the laboratory, provide a faster result (15 min) and can be produced much faster, cheaper and in large quantities, allowing for swift upscaling of testing capacity. They can be highly specific, but are generally not as sensitive as molecular tests, making them effective for identifying infected persons displaying high virus shedding, who are hence most infectious [4], [5].

Although laboratory-based NAAT is still considered to be the reference standard for SARS-CoV-2 diagnosis, the WHO recommends the use of Ag-RDTs as a decentralizable, faster and reliable option, provided that the tests meet the WHO standards for Ag-RDTs (≥ 80% sensitivity and ≥ 97% specificity among symptomatic individuals) [6], [7].

The continuous evolution of SARS-CoV-2 has already resulted in the emergence of several variants of interest (VOI) and of concern (VOC) which can be associated with increased transmissibility and/or immune escape [8], [9]. Therefore, genomic surveillance to allow early identification, detection, monitoring and reporting of emerging variants can be considered to be equally important as epidemiological surveillance in achieving effective mitigation and containment of the virus. Moreover, virus whole genome sequences can be used to investigate spatiotemporal spread and transmission routes, and can help in the design of diagnostic assays, antivirals and vaccines [10]–[12].

Genomic surveillance of SARS-CoV-2 can be achieved by complete genome sequencing (the golden standard) or by detection of spike protein mutations that are indicative for specific variants through sequencing of a limited region of interest within the receptor binding domain (RBD) of the viral genome [13], [14]. These techniques require the retrieval of viral genetic material from a patient sample, classically the leftover of a nasopharyngeal sample used for NAAT.

Based on our previous experience with the preservation and transport of viral material on paper strips [15]–[18], we wanted to test whether the leftover viral material in the cellulose carrier in the test strips used in SARS-CoV-2 Ag-RDTs is sufficient to allow extraction of intact viral nucleic acids and subsequent RT-qPCR, Sanger sequencing and/or whole genome sequencing (WGS).

Several other viruses are also frequently diagnosed with Ag-RDTs, and also in these cases obtaining supplementary information regarding the type or variant through sequencing can be of importance. Genomic surveillance of influenza viruses is being performed by labs worldwide within the framework of the WHO’s Global Influenza Surveillance and Response System (GISRS). They provide essential information regarding efficacy of vaccines and antiviral drugs against currently circulating influenza viruses, vaccine strain selection and the potential spill-over of animal influenza viruses to humans. The surveillance of circulating genotypes of rotavirus is of crucial importance for detection of novel emerging genotypes and/or antigenic drift of strains that can occur through vaccination and can lead to decreased efficacy or failure of vaccines. For the cases of hepatitis of unknown etiology among young children that were under investigation by ECDC at the time of writing [19], an association with a (novel variant of) human adenovirus 41 (Adv41) is being considered [20], [21]. Typing of positive adenovirus samples can therefore be of clinical and epidemiological relevance.

In this study, we evaluate the possibility to retrieve leftover material of several viruses from a range of Ag-RDTs, and after longterm storage at room temperature, for use in molecular genetic analysis.

## MATERIALS AND METHODS

### Samples

For an optimal result, Ag tests need to be performed with fresh samples containing live virus. Since it was not feasible to perform all Ag tests immediately on infected patients, with sampling using the swab provided by the supplier, we used anonymized stored nasopharyngeal patient samples as a proxy. For use on an Ag-RDT, samples need to be stored in PBS and not in inactivated medium such as zymo, inactive blue or any other virus-inactivating medium that denatures proteins, since Ag-RDTs detect intact viral proteins.

Our laboratory functions as the Belgian national reference center (NRC) for coronaviruses, adenoviruses and rotaviruses. The positive samples and positive Ag-RDTs used in this study were anonymized leftover materials of diagnostic samples, tested by the NRC. The use of patients’ residual materials by the NRC was approved by the UZ Leuven Ethics Committee.

### Rapid Ag tests (Ag-RDTs)

SARS-CoV-2 Ag-RDTs that were used in this study are: Roche SARS-CoV-2 Rapid Antigen Test Nasal, Roche Diagnostics, Basel, Switzerland, Ref. 9901-NCOV-01G and 9901-NCOV-06G (**Roche nasal**); FlowFlex SARS-CoV-2 Antigen Rapid Test (Self-Testing), Acon Laboratories Inc., San Diego, CA, USA, Ref. L031-118P5 (**FlowFlex**); Newgene COVID-19 Antigen Detection Kit – Nasal Swab, Newgene Bioengeneering, Hangzhou, China, Ref. COVID-19-NG21 (**Newgene**); Coris COVID-19 Ag K-SeT, Coris BioConcept, Gembloux, Belgium, Ref. K-1525 (**Coris COVID-19**); Alltest: nasal swab test, Hangzhou Alltest Biotech Co. Ltd., Hangzhou, China, Ref. INCP-502-N (**Alltest**); Boson Rapid SARS-CoV-2 Antigen Test Card, Xiamen Boson Biotech Co., Ltd., Xiamen, China, Ref. 1N40C5-2 (**Boson**). SARS-CoV-2 / Influenza dual Ag tests that were used are: Orient Gene Influenza & COVID-19 Ag Combo Rapid Test Cassette, Zhejiang Orient Gene Biotech Co., Ltd, Zhejiang, China, Ref. GCFC-525a (**Orient**); AMP Rapid Test CoV-2 Ag + Flu A+B, AMEDA Labordiagnostik GmbH, Graz, Austria, Ref. RT2962 (**AMP**); Nadal COVID-19 Ag+Influenza A/B plus test, Nal von minden GmbH, Moers, Germany, Ref. 243204N-20 (**Nadal**). For Adenovirus 40/41 and Rotavirus dual Ag test we used the Coris GastroVir K-SeT, Coris BioConcept, Gembloux, Belgium, Ref. K-1516 (**Coris Gastro**). Tests are referred in Table 1 as indicated in bold after every test.

**Table 1:**
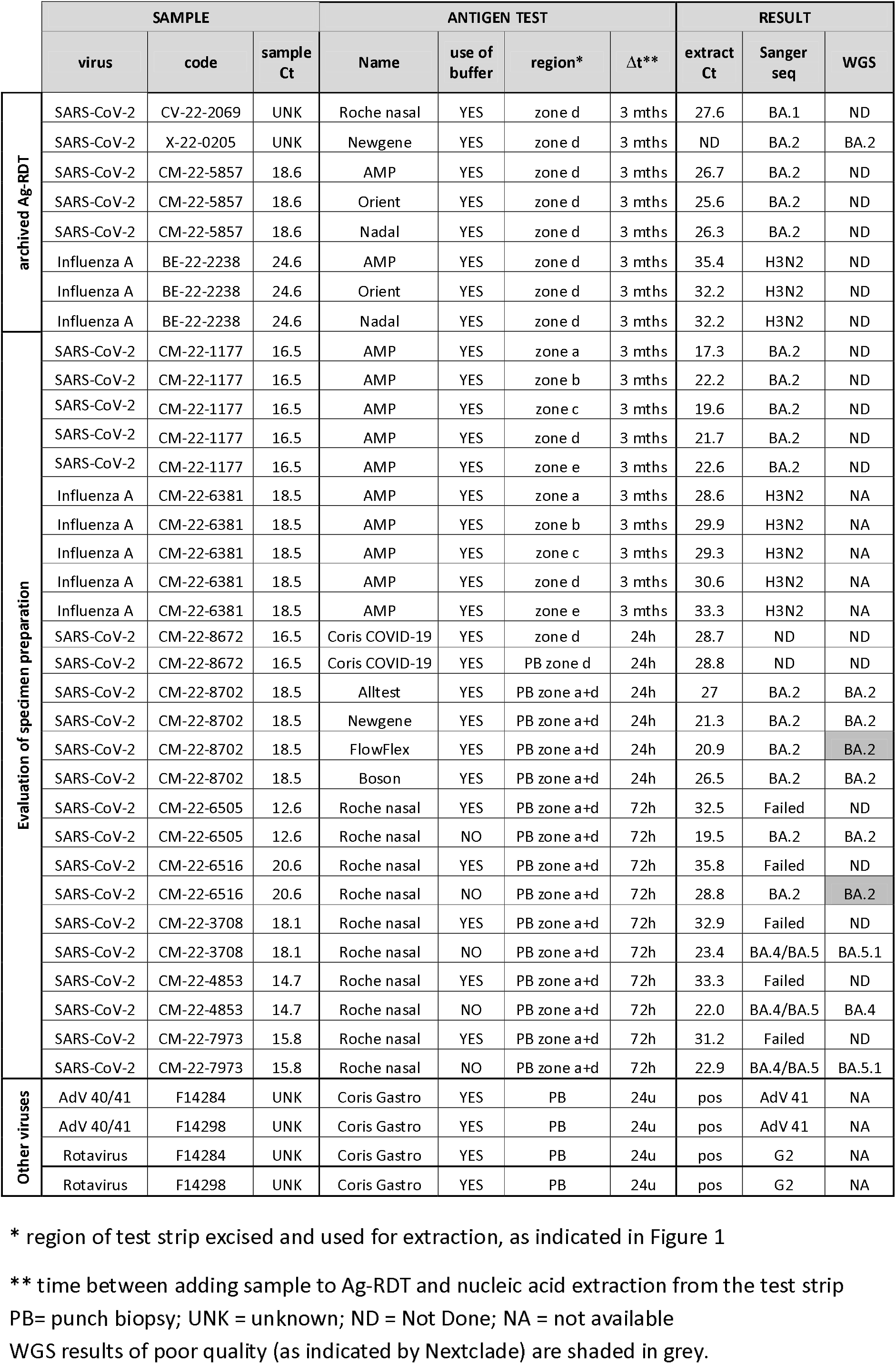
Retrieval of viral material from rapid Ag tests.

Direct testing of patients with the Ag-RDTs was done as described in the test instruction manual. For testing of different Ag-RDTs with positive samples of known viral loads, nasopharyngeal swabs in PBS that had been stored at 4°C were used. Hundred μl of the PBS solution was mixed with the buffer included in the test kit. The Ag-RDT test was further performed as described in the test instructions manual. To assess the effect of the Ag-RDT test buffer on nucleic acid yield, we also performed Ag-RDT tests without using the buffer solution, by transferring 100 μL of PBS solution directly on to the sample window of de test cassette. Ag-RDTs run with PBS without patient sample were used as negative controls.

### Nucleic acid extraction from Ag-RDT test strips

Ag-RDT cassettes were opened by removing the cover, the test strip was taken out and different parts of the test strip were excised with scissors as indicated in Fig. 1.

**Figure 1:**
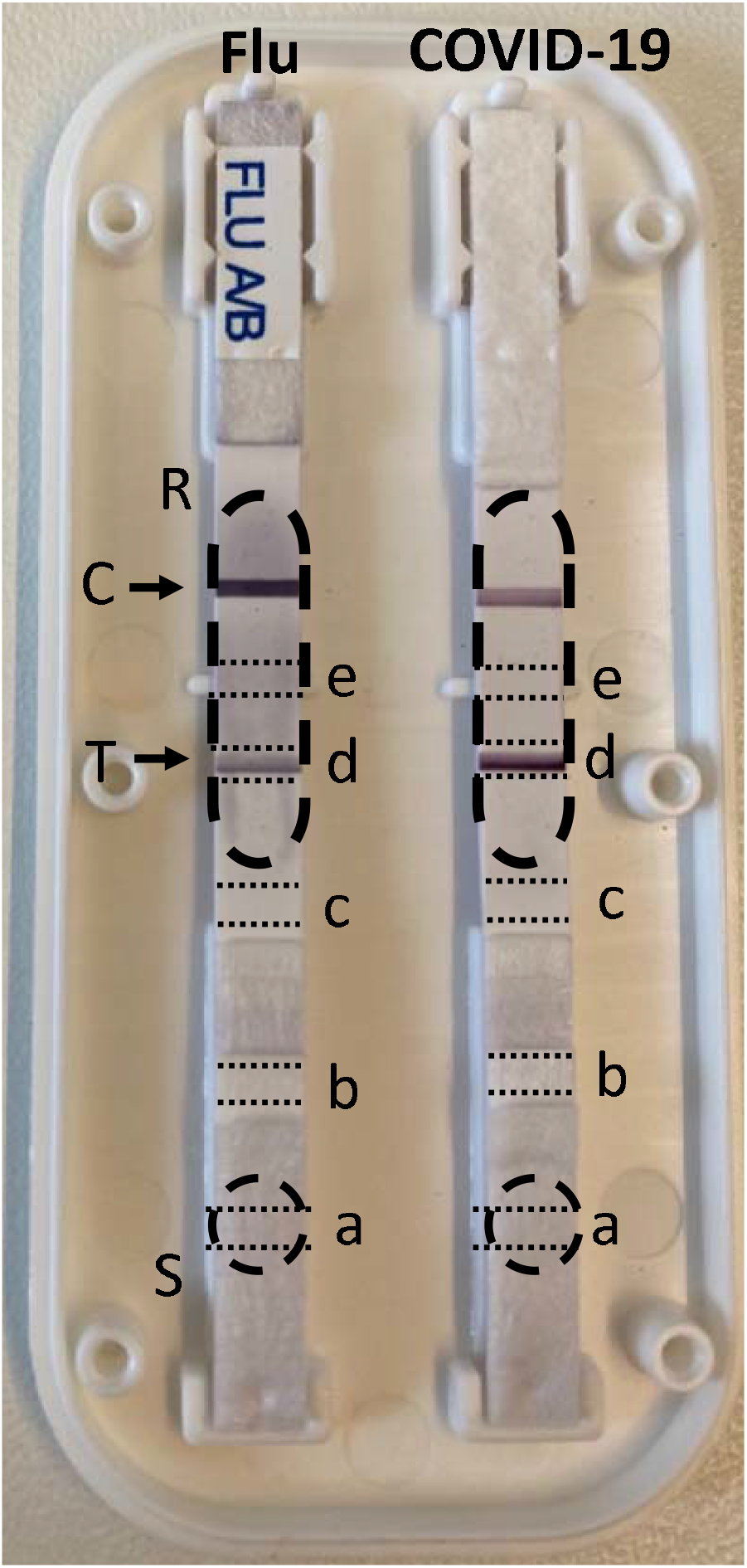
Opened cassette of an Orient Gene Influenza & COVID-19 Ag Combo Rapid Test, with dashed lines indicating the position of the sample windows (S) and result windows (R), displaying a positive test line (T) and control line (C). Regions excised for nucleic acid extraction (a-e) are delineated with dotted lines.

Punch biopsies were taken from unopened Ag-RDT cassettes at the zone of the sample window and at the zone of the positive test line, by using a 2 mm diameter sterile disposable biopsy punch (KAI medical disposable biopsy punch Ref. BP-20F). The biopsy or excised part of the Ag-RDT test strip was directly added to 275 μL lysis buffer in a 96 well lysis plate for extraction with the MagMAX Viral/Pathogen II Nucleic Acid Isolation Kit (ThermoFisher Scientific), according to the protocol for KingFisher Flex.

### Detection of SARS-CoV-2 and Influenza with RT-qPCR

To detect SARS-CoV-2, an RT-qPCR targeting a region in the N2 gene was performed on the QuantStudio™ 7 Flex Real Time PCR system (Applied Biosystems, ThermoFisher). To amplify the N gene region, a reaction mix was made using 5 μL TaqMan™ Fast Virus 1-step Master Mix (Applied Biosystems, Cat 4444434), 1.5 μL primer/probe mix from the 2019-nCoV CDC EUA kit (IDT, Cat 10006606) supplemented with 8.5 μL RNase free water to a total volume of 15 μL.

For Influenza qPCR a reaction mix was made using 5 μL TaqMan™ Fast Virus 1-step Master Mix (Applied Biosystems, Cat 4444434) with 0.12 μL forward primer (INFA-3) 5’TCTCATGGAATGGCTAAAGACAAG-3’ (50μM) together with 0.2 μL probe (INFA-FP) FAM-5’- TTCACGCTCACCGTGC-3’-MGB (20μM) and 0.12 μL reverse primer (INFA-2) 5’- CAAAGCGTCTACGCTGCAGT-3’ (50μM) supplemented with 9.56 μL RNase free water to a total volume of 15 μL.

Five μL of viral RNA was added to the reaction mixes. Thermal cycling conditions were 5 minutes at 50°C, 2 minutes at 95°C, followed by 45 cycles of 3 seconds at 95°C and 30 seconds at 60°C. Analysis was done using the QuantStudio Real-Time PCR Software (Applied Biosystems, ThermoFisher).

### Sanger sequencing for virus typing

Sanger sequencing was performed as described in Bloemen et al., 2022 with SARS-CoV-2 primers (S-VOC-F/S-VOC-R). Additional primers for partial sequencing of influenzavirus (AM_FW151/AM_RV397) were described by Schweiger et al., for adenovirus (F-hex1deg/R-hex2deg) by Allard et al. and for rotavirus (F-BEG9/R-END9) by Gouvea et al. [22]–[24].

### Complete genomic sequencing of SARS-CoV-2

Complete genome sequencing of SARS-CoV-2 on RNA extracted from the Ag-RDT was done with the nanopore technique using the ARTIC protocol as described in Wawina-Bokalanga et al. [25]. Sequences were analyzed with Nexclade v 2.0.0 (https://clades.nextstrain.org)

## RESULTS

### Sequencing of SARS-CoV-2 from archived Ag-RDT

A positive SARS-CoV-2 Ag-RDT (SARS-CoV-2 Rapid Antigen Test Nasal, Roche, Ref. 9901-NCOV-06G), that had been used according to the test kit instructions to test an infected patient and was kept for documentation purposes, had been stored at room temperature and not shaded from sunlight for 3 months. As a proof of concept, we tested whether residual viral genetic material could be extracted from the test strip. On this strip, the positive test line was still clearly visible, and extraction was done on the positive test strip zone of the test (zone d, indicated in Fig. 1). SARS-CoV-2 RNA could still be detected by RT-q-PCR, with a Ct value of 27.6. Sanger sequencing on the extracted material allowed to unambiguously determine the virus variant as Omicron BA.1, based on the partial sequence of the S-gene RBD (Table 1).

We subsequently applied the same method to a Newgene SARS-CoV-2 Ag RDT that had been run with leftover material of a positive nasopharyngeal swab sample, and was stored under the same conditions, and were able to perform complete viral typing by WGS. Likewise, viral material could be retrieved and sequenced from archived SARS-CoV-2/influenza dual Ag-RDTs that had been run with positive nasopharyngeal swab samples (Table 1).

### Evaluation of specimen preparation

The lateral flow rapid Ag tests used in this study all contain a test strip on which the sample is added via the test window. The sample is absorbed onto the sample pad of the test strip, and is transported via capillary flow to the distal end of the strip, flowing over the conjugate pad, test line and control line. To investigate in which area of the test strip the largest amount of viral material from the sample is maintained, and is thus best suited to be used to extract viral material, different zones of the test strip were excised with scissors and used for nucleic acid extraction and RT-qPCR (Fig. 1). A dual antigen test for influenza and COVID-19 (AMP Rapid Test CoV-2 Ag + Flu A+B) that had been run with an influenza positive sample and a SARS-CoV-2 positive sample, was used for nucleic acid extraction 3 months after the Ag test was run. The viral nucleic acid yield at different positions was determined by RT-qPCR for influenza A and SARS-CoV-2 respectively (Table 1: CM-22-1177, CM-22-6381). At all positions that were tested, viral genomic material could be harvested, with the sample pad resulting in the highest yield for both SARS-CoV-2 and influenza, and the zone distal from the test line giving the lowest yield for both viruses. We were able to successfully perform Sanger sequencing on all extracts.

To evaluate the possibility of performing extraction from Ag-RDTs in a high throughput environment, we also tested the use of a punch biopsy sample taken at the test line of the strip. These punch biopsies can be picked without opening the test cassette. The use of a punch biopsy versus excision of the test line zone of the strip was compared using a Coris COVID-19 Ag test, and the resulting Ct value were approximately the same (Table 1: CM-22-8672). Since the biopsy procedure can be standardized and is relatively easy to use, we decided to use punch biopsies in further experiments.

To evaluate whether viral material is conserved at a comparable degree in different brands of SARS-CoV-2 Ag-RDTs, tests that were commonly used in Belgium at the time of this investigation were run in parallel using the same positive sample (Ct of 18.5). This resulted in a clear positive test line on all tests that were analyzed. Twenty-four hours after performing the Ag test, a punch biopsy was taken from the test line zone and the sample pad of the test strip and used for extraction, followed by RT-qPCR, Sanger sequencing and WGS. All brands of SARS-CoV-2 Ag-RDTs that were tested allowed detection of SARS-CoV-2 by RT-qPCR in the punch biopsy, and in all cases variant typing by Sanger sequencing was possible (Table 1: CM-22-8702). This indicates that conservation of viral material in a used Ag-RDT did not only occur in one specific brand, and that SARS-CoV-2 Ag-RDTs in general can be used for genomic surveillance. We did however notice quite extensive differences in the RNA yield that could be obtained, as indicated by the SARS-CoV-2 Ct value after extraction. Samples extracted from Alltest, Newgene and Boson resulted in WGS data of good quality whereas the FlowFlex sample was typable by WGS but data quality was poor.

Since the Ag-RDTs use buffers with different compositions, the effect of the buffer on RNA yield was tested. Samples were loaded on an Ag test with and without addition of the test kit buffer (Table 1,test performed on Roche nasal). For all samples tested, the Ct value of the extract was around 10 points higher when buffer was used, which equals approximately a 1000-fold lower concentration of viral genomic material. In general, the results of Sanger sequencing were successful when no buffer was used and good data quality was obtained by WGS. This indicates that some buffer components are deleterious for the viral genetic material, resulting in lower RNA concentrations and interfering with sequencing. WGS was not performed on samples that failed for Sanger sequencing given the high cost of nanopore sequencing.

### Retrieval of viral material of influenzavirus, adenovirus and rotavirus from Ag-RDTs

An influenza A virus positive sample (nasopharyngeal swab in viral transport medium), that was tested in the routine diagnostic lab of the NRC, had been run on the influenza test strip of the AMP, Orient and Nadal SARS-CoV-2/influenzavirus dual antigen tests. A positive test line for influenza A was obtained with all three Ag tests. Three months after performing the Ag test, the test line zone was excised with scissors, and viral material was extracted. After extraction, sequences of influenza A could be retrieved by Sanger sequencing of part of the M gene for all three Ag tests.

Two fec al samples that were positive for both adenovirus and rotavirus by a routine diagnostic test in the NRC lab were run on Coris GastroVir K-SeT rapid Ag tests. One sample displayed a positive test line for both adeno- and rotavirus on the strip, while the other sample displayed a positive test line for rotavirus but not for adenovirus. Punch biopsies were taken from the sampling pad, the rota test line and the adeno test line of the strips. After extraction of these biopsies, PCR and Sanger sequencing in the VP7 gene was performed for rotavirus detection. For both samples we detected the presence of rotavirus G2 in extractions of the biopsies taken at all 3 sampling sites of the test strip. The adenovirus PCR was also positive for both samples, and sequences of adenovirus 41 were retrieved, again at all 3 sampling sites of the test strip, also in the sample that did not display a positive test band.

## DISCUSSION

Whereas RT-qPCR continues to be the gold standard for COVID-19 diagnosis, rapid Ag-RDTs are widely used since they offer the benefit of a very short time to results, are cheaper, and easy to use. A disadvantage of testing through Ag-RDTs so far was that no leftover sample material was available for genomic characterization of positive samples. Our finding that the test strip used in rapid Ag tests are suited to preserve intact viral genomic material, even for months, and can serve as source material for genomic characterization of the virus, could be a gamechanger in the COVID-19 testing strategy.

We demonstrated that a SARS-CoV-2 Ag-RDT that was stored at room temperature up to 3 months could still serve as source material for genomic surveillance. This implies that used Ag test can be easily sent to sequencing facilities, even by regular mail. This provides an easy and cheap way to establish baseline genomic surveillance in countries/regions with limited resources, which would otherwise remain blind spots thus allowing for novel virus variants to remain under the radar. This could therefore mean an important improvement in the global coverage of genomic surveillance.

In this study, we have tested a total of 9 different Ag-RDTs for SARS-CoV-2 detection (with or without concurrent influenza detection), and were able to successfully perform Sanger sequencing on extracted material from all of these. The difference between the original Ct in the sample and the Ct value after extraction of the test strip largely differed depending on the Ag-RDT used. Our results indicated that the buffer used by some Ag tests contains a chemical compound that breaks down the viral RNA and has a negative effect on the quality of sequencing data. For instance, in case of the Roche nasal test, use of the test kit buffer resulted in a much lower viral RNA yield and made the extract unuasable for sequencing. Use of the same Ag test when putting the samples directly on the cassette (without buffer) resulted in much better yields and good sequencing results, both by Sanger and by WGS.

Nazario-Toole et al. demonstrated recently that whole genome sequencing of SARS-CoV-2 from excess clinical specimens processed using the BinaxNOW^™^ COVID-19 Ag Card was feasible [26]. They however did not assess the effect of prolonged room temperature storage of completed Ag cards, nor did they study different brands of SARS-CoV-2 Ag-RDTs, or Ag-RDTs for other viruses.

We have shown that this method is more widely useable, by demonstrating that genomic material of influenza virus, adenovirus and rotavirus can also be retrieved from rapid Ag tests. Especially during the upcoming winter seasons, when an upsurge in COVID-19 is to be expected with concomitant circulation of other respiratory pathogens such as influenza virus, the use of Ag-RDTs that allow combined detection of multiple respiratory pathogens, such as COVID-Flu a/b combo tests, can prove to be a very cost-efficient testing strategy. We have now shown that the use of these Ag tests can be combined with genomic surveillance, which will remain equally important in the monitoring of novel variants.

We were able to confirm the usability for 10 commercial Ag-RDTs which are commonly used in Belgium. Since it was not possible to test all commercial Ag-RDTs, a validation experiment for each brand and with use of different buffer compositions should be performed prior to large scale implementation of a certain Ag-RDT in genomic surveillance. We plan to further optimize the method for high-throughput processing.

## Data Availability

All data produced in the present study are available upon reasonable request to the authors

## ACKNOWLEDGEMENTS

UZ Leuven, as national reference centre, is supported by Sciensano, which is gratefully acknowledged.

